# Effectiveness of using reward strategies combined with transcranial direct current stimulation (RstDCS) on motivation in chronic stroke patients with upper limb disorders: study protocol for a randomized controlled trial

**DOI:** 10.1101/2023.03.22.23287541

**Authors:** Ping Zhou, Wenxi Li, Jingwang Zhao, Siyun Chen, Yufeng Chen, Xia Shen, Dongsheng Xu

## Abstract

**Introduction:** Stroke survivors tend to have low motivation for rehabilitation, which prevent them from completing the rehabilitation training task effectively and participating in daily activities. Reward strategies can stimulate patients’ motivation for rehabilitation, but the duration of their effect is unknown. tDCS is a technique capable of promoting plastic changes and functional reorganization of cortical functional areas. When it acts on dlPFC, it can improve the functional connectivity of brain areas associated with goal-directed behavior. Reward strategies combined with tDCS (RstDCS) have been shown to make normal individuals work harder at performing tasks. However, there are no studies on whether this combined strategy improves motivation in stroke patients and whether it has a long-lasting effect on motivation maintenance.

**Methods and analysis:** 87 stroke survivors who have low motivation and upper extremity dysfunction will be randomized to receive conventional, Rs, or RstDCS treatment. Patients in each group will undertake 15 sessions of tDCS real or sham stimulation of the left dlPFC for 3 weeks in conjunction with conventional rehabilitation or reward strategy training. Following this, the RS treatment group and RstDCS treatment group will be given a 3-week outpatient self-monitoring training program, while the conventional group will be given home rehabilitation instruction prior to discharge.

Rehabilitation motivation is the primary outcome measure and will be assessed using RMS. RMS,FMA, FIM and ICF activity and social engagement scale will be compared at baseline, 3 weeks, 6 weeks, and 3 months after enrollment.

**Ethics and dissemination:** The study has been approved by the Ethics Committee of Yueyang Hospital of Integrated Chinese and Western Medicine Affiliated to SHUTCM (2021-122). All participating patients will provide written informed consent. The results will be disseminated through (open access) peer-reviewed publications, networks of scientists, professionals, and the public.

**Trial registration number:** ChiCTR2300069068

## Article summary

### Strengths and limitations of this study

1. We adopt simple and feasible reward strategies, coordinated with neural regulation technology, which are intrinsic stimulation and extrinsic enhancement, to provide the industry insider with the preliminary exploration approach of a comprehensive strategy to improve the patients’ rehabilitation motivation.
2. We used a variety of methods, such as behavioral observation and others’ ratings, to observe patients’ rehablitaton motivation.
3. In this study, although behavioral observation and other rating scales were used to evaluate patients’ motivation, there was a lack of assessment tools that could evaluate patients’ motivation status in real-time.
4. Whether the benefits of rehabilitation motivation enhanced by RstDCS can reduce the burden and cost of time and money for patients and their families need to be verified with a larger sample size.

## Introduction

50% of stroke survivors have upper limb (UL) functional impairment, which seriously influences daily life^1^. Fortunately, the recovery period of upper limb function is long and there is still the possibility of improvement even after 6 months^2^. Evidence suggests a positive correlation between a therapeutic dose and functional recovery^3^. Therefore, it is necessary to encourage patients to exercise for a longer time and increase the number of exercise repetitions^4^. However, the shortage of rehabilitation resources and the high cost of medical treatment mean that most patients do not receive the amount of treatment they should receive under the supervision of therapists during their brief stay in the hospital, let alone the time to return home ^5^. The key to solve this problem is for patients to have positive motivation for rehabilitation in order to improve the efficiency of rehabilitation training so that rehabilitation services can be more sufficiently utilized^6^. Therefore, more and more researchers began to study how to enable patients to take active rehabilitation exercises and increase exercise dose without professional supervision (such as non-treatment time in the hospital and time back home)^7^.

The most important cause of low motivation in stroke patients is post-stroke apathy^8,9^. It is present in about 1/3 of stroke patients, with symptoms beginning as early as 4 days after stroke^10^. It remains in most post-stroke patients up to one year after stroke, with only a minority improving^11^. Apathy is manifested in the gradual decline of rehabilitation motivation and the reduction or refusal of the participation in exercises, resulting in a corresponding decrease in exercise dose^12,13^. If the patient’s rehabilitation motivation continues to be low, then no amount of medical resources are available. On the contrary, when the rehabilitation motivation is improved, patients can not only make useful judgments about themselves, actively carry out rehabilitation training, have the confidence to face difficulties positively, but also enhance motor relearning^14–17^. Six months after stroke, not only psychogenic apathy may be caused by psychological and social factors, but also organic secondary apathy may be caused by neurobiological changes^18^. The possible neural mechanism is the impairment of the brain network related to goal directed behavior (GDB)^19^, which includes the anterior Cingulate cortex (ACC) and the nucleus accumbens (NAcc)^8,20^. Another likely reason is that infarction may lead to functional disruption of the connective area after an acute stroke^21^.

At present, the mainly purpose of improving patients’ motivation is promoting their decision-making ability and motor relearning ability. Among them, the methods to promote the decision-making ability of patients include motivational interviewing^22,23^ integrated approach to individual rehabilitation programs based on goal orientation and self-management^24–27^ interventions developed from behavioral economics ^28,29^. Such approaches are based on social cognition theory^30^ and self-determination theory ^31^. The process of education, coaching and co-setting goals can improve patients’ self-efficacy and promote their internal motivation for recovery^32^. Methods to improve patients’ motivation and thus motor relearning include sensor technology ^33,34^, robot-assisted technology and virtual reality equipment^35,36^. Such methods promote patients’ motor motivation by providing refined feedback, and as a form of intrinsic reward, enhance patients’ ability to analyze and process feedback information, thus improving patients’ motor accuracy, proficiency, speed and coordination^37^. However, less attention has been paid to the fact that the activation of reward systems associated with motivation and motor learning is reduced in the brains of stroke patients compared to normal subjects, so that even with relevant strategies, the effect of improving motivation is not significant^13,38^. Stronger stimulation or treatment is needed to normalize reward processing^39^. Non-invasive neuromodulation techniques such as transcranial direct current stimulation (tDCS) and transcranial magnetic stimulation (TMS) can normalize the activation of motivation-related brain regions in stroke patients ^38,40^. Some evidence suggests that TMS or tDCS can improve apathy and alter the balance of activity in prefrontal cortex and subcortical areas to promote successful self-regulation in patients^41,42^.

Although there are many interventions to promote patients’ motivation to recovery, improvements are needed. First, most approaches focus only on patients’ motivation during treatment; maintenance of motivation after treatment is rarely mentioned^43^. However, the maintenance of high motivation is most notable in patients with chronic disease, which is directly related to whether the patient initiates exercise without supervision. Second, the cost-effectiveness, health care resource consumption, treatment frequency, availability, acceptability, and generalizability of all current motivational interventions need to be investigated and improved. Third, there is a lack of interventions to improve the treatment of secondary alterations in stroke, such as functional decline in brain areas associated with motivation and its associated dysfunctional brain network connections. The way forward should be to combine motivational interventions that arise from multidisciplinary interactions. Last but not least, there is no comprehensive approach to assessing motivation in stroke patients, and a standardized process for assessing motivation for rehabilitation should be established.

Based on the above background, the group designed a simple and feasible treatment plan using less medical resources and costs, that is a reward strategy combined with a neuromodulation treatment plan, hoping to modulate patients’ neural activity through tDCS and enhance the functional connectivity of brain regions associated with goal-directed behavior. In addition, we should work with patients to set treatment goals and give them external rewards to affirm their autonomy and self-drive to complete the exercise, thus stimulating their intrinsic motivation and maintaining the initiative of the rehabilitation program from the whole process of "hospital, family, and society".

## Objectives

This randomized controlled trial aims to investigate the effectiveness of rewarding strategy combined with tDCS in improving rehabilitation motivation compared to rewarding strategy group without tDCS and conventional treatment group in 87 patients with chronic stroke (onset over 6 months). With this pilot study, we hope to verify whether the reward strategies designed to increase rehabilitation motivation can provide a feasible approach to the entire rehabilitation process in "hospital, family and society". The primary objective of this study is to compare the scores of rehabilitation motivation scale and times patients participating in doing exercises without supervision at baseline, 3 weeks, 6 weeks after treatment and 3 months follow-up. The secondary objective is to evaluate the structure and function, activities and participation of patients within the framework of ICF to observe whether rewarding strategy can improve rehabilitation motivation, motor ability, daily living ability and quality of life through self-report and assessment instruments.

## Methods and analysis

### Study design

The reward strategy combined with tDCS treatment(RStDCS) for improvement of rehabilitation motivation after stroke study is a randomized, single-blind, controlled clinical trial. Subjects will be randomly assigned to reward strategy combined with tDCS treatment(RStDCS), rewarding strategy with sham tDCS treatment (RS group), conventional treatment with sham tDCS treatment and then treated with upper extremity rehabilitation. This is a single-blind study. All patients receive either true or sham tDCS stimulation and are given an exercise program manual (content varies with different requirements). And the evaluators and statisticians do not know what group the subjects belongs to. During the trial, the subjects will be assessed at 0, 3, 6 weeks and followed up at 3 months by means of assessment scales and behavioral observations. Prior to the assessment, the evaluator will be trained. Assessment scales include RMS (Rehabilitation Motivation Scale), FMA (Fugl-Meyer Assessment Scale), FIM (Function Independent Measure), ICF (ICF Activity and Participation Scale). Table 1 shows the schedule of enrolment, interventions, and assessments.

**Table 1:**
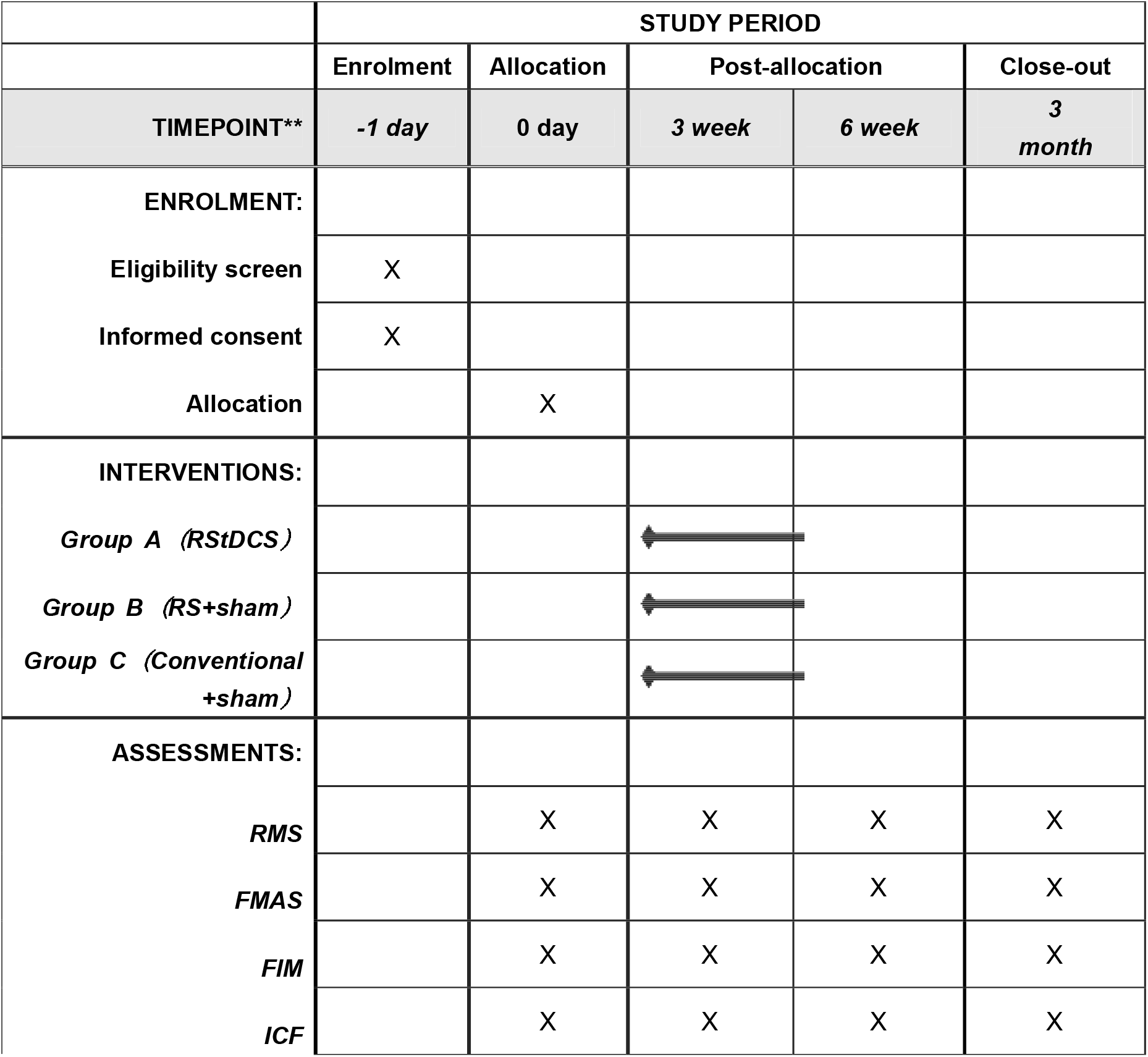
The schedule of enrolment, interventions, and assessments

### Study population and recruitment

Initial steps are described in Figure 1. Participants (total of 87) are recruited from the inpatient and the outpatient department of Rehabilitation Medicine Center of Yueyang Hospital of Integrated Traditional Chinese and Western Medicine affiliated to Shanghai University of Traditional Chinese Medicine. Patients with previous confirmed upper limb or hand dysfunction after stroke will be screened for eligibility to participate in this study based on the criteria. After the patient has been assessed as eligible by the rehabilitation physician, he/she will receive initial study information. After at least 2 weeks of reflection, patients are invited to meet with the research physician to discuss any remaining questions and sign the informed consent.

**Figure 1:**
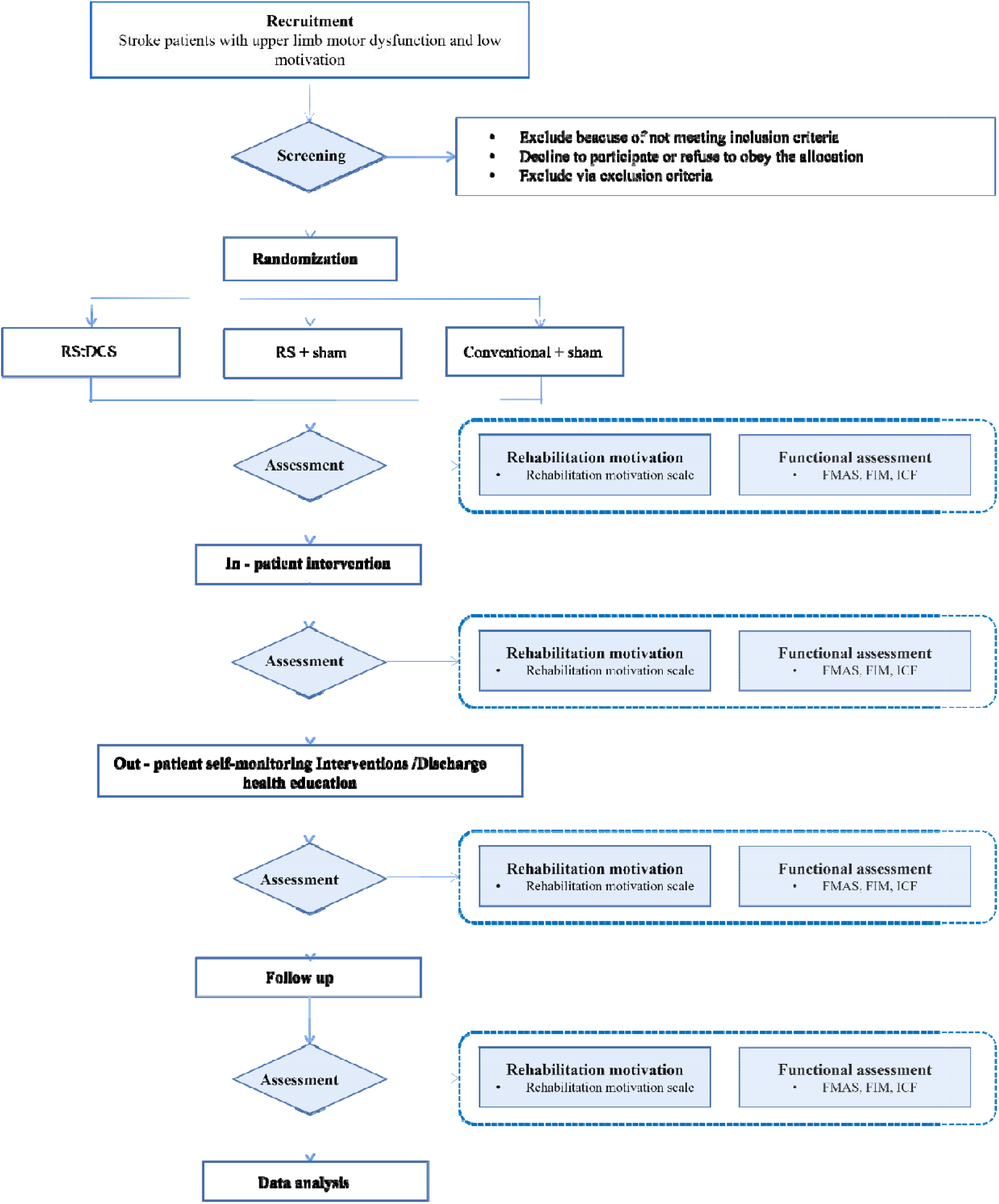
Flowchart of study design

### Inclusion and exclusion criteria

Patients should meet the following criteria to be eligible for the study:

1. The patient provides written informed consent, is able to understand the content of the study, understands the requirements for follow-up visits, and is willing to complete the questionnaires and provide the required information at follow-up visits;
2. Patients diagnosed as stroke met the diagnostic criteria of ischemic stroke set by the National Cerebrovascular Academic Conference in 2014;
3. Age ≥ 18 and ≤70 years old, no gender limitation, right-handed;
4. No cognitive impairment or communication impairment, Mini-Mental State Test (MMSE) score of ≥ 27;
5. First onset and the course of disease is more than 6 months;
6. Upper limb and hand dysfunction, Brunnstrom Stage ranged from I to VI;
7. Subjects’ rehabilitation motivation Scale score of ≤25.

The exclusion criteria of this study are:

1. Patients with serious systemic diseases such as cardiopulmonary diseases who cannot tolerate rehabilitation treatment;
2. (History of) Psychosis, major depression (suicidal tendency) or epilepsy;
3. Serious systemic diseases such as diabetes and uremia exist;
4. Severe joint contracture;
5. Suffering from any disturbance of consciousness caused by any cause;
6. Contraindications for TMS/tDCS and fMRI examination according to safety guidelines, such as metal foreign bodies or other implanted electronic devices;
7. Auditory or visual impairment may affect assessment and treatment;
8. Use of drugs that change the excitability of cerebral cortex (antiepileptic drugs, sedatives and hypnotics, etc.);
9. Obvious pain, sleep disturbance, mental disorder.

### Randomization and allocation

A random number table will be used for randomization to divide patients into three groups. Allocation is concealed with the use of serially numbered, opaque, sealed envelopes. When the researcher determines the eligibility of the subjects, the envelopes are opened in sequence and the subjects are assigned to the corresponding test group.

The researcher who generates and preserves the randomly assigned sequences are not involved in the trial. After other researchers determine the eligibility of the subjects, they are placed into different groups according to the order in the envelope. The trial groups will not be known to the subjects and the therapists who provide them with routine care.

### Interventions

Figure 2 shows the groups overview and research implementation process. The control groups receive conventional upper limb rehabilitation training without rewarding strategies and stimulation of tDCS. The participants of control group will be trained for 20min per day, 5 days a week for 3 weeks while staying in a participating rehabilitation hospital. The control groups also get rehabilitation education after being discharged. The RStDCS group will receive upper limb rehabilitation training combined with rewarding strategies and non-invasive neuromodulation techniques, which is to stimulate the intrinsic motivation of patients by using tDCS to stimulate the left dorsolateral prefrontal cortex (dlPFC) where is thought to be an important role in computing the predicted capacity to successfully exert effort, enabling the dorsal anterior cingulate cortex (dACC) to compare the required work with the potential benefits^44^. The anodal electrode is positioned over left dlPFC (between F3 and F7) according to the International 10 – 20 system. Cathode electrode was placed contralaterally to the anode on right shoulders. During the anodal stimulation condition, a continuous current (1.5 mA) was delivered (35 cm^2^ electrodes) for a current density of 0.043 mA/cm^2^. Treatment is given once a day, 20 minutes for each time, a total of 15 times in 3 weeks.

**Figure 2:**
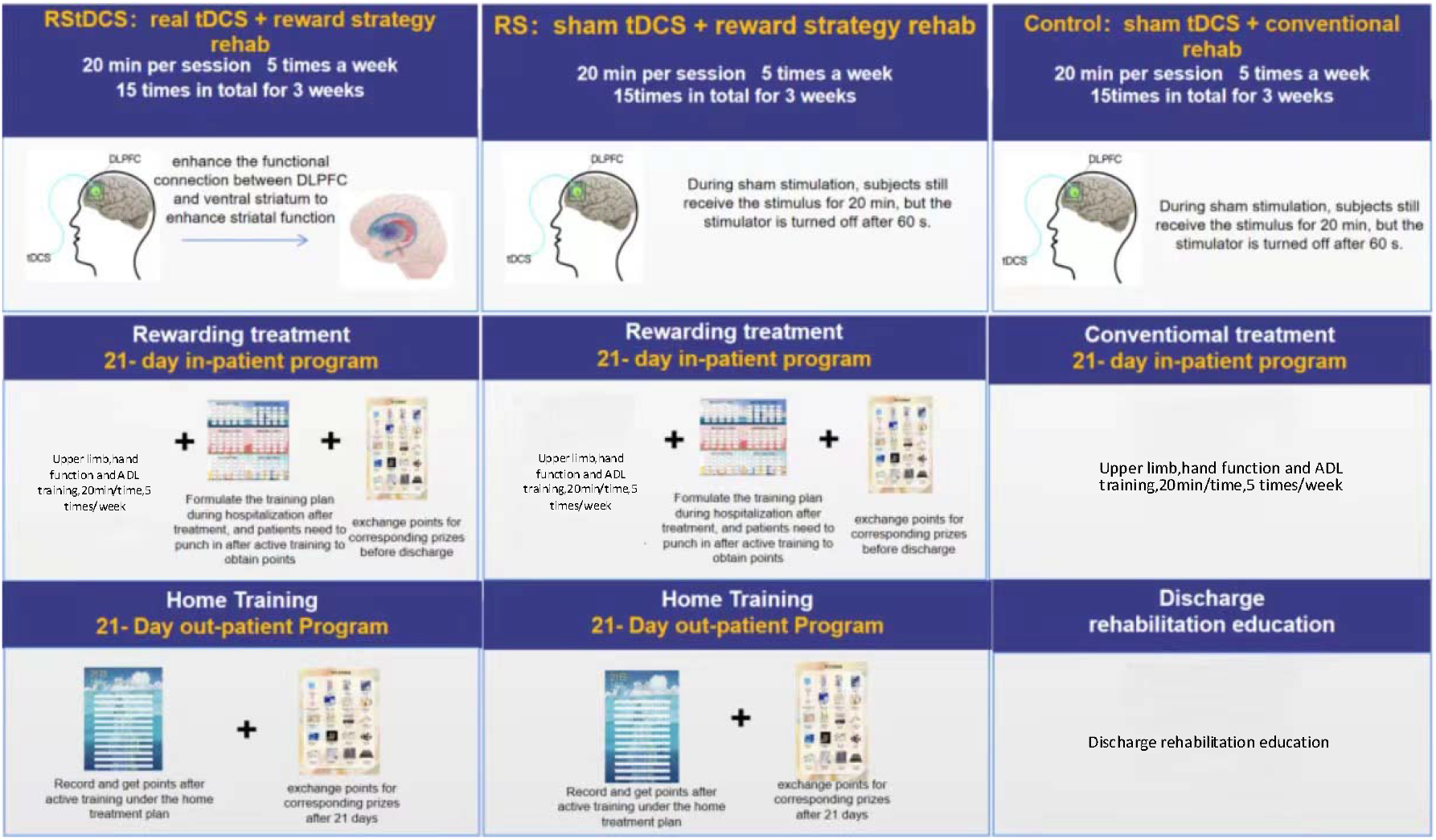
Groups overview and research implementation process

Compared to the RStDCS group, the RS group will receive upper extremity rehabilitation based on reward strategies, in conjunction with sham tDCS stimulation in the same position of the RStDCS group. After 60s of stimulation, the patient’s stimulator will be automatically turned off.

The rewarding strategies mean giving them extrinsic rewards based on the recognition of their independent performance and ability to observe whether it can maintain the active rehabilitation motivation of patients from the whole process of "hospital-home-society". Specifically, a 21-day inpatient training task manual is developed for patients with chronic stroke with hand dysfunction and low motivation during hospitalization, and a 21-day home training program is customized for patients returning home. Figure 3 shows the schematic diagram of reward strategy self-monitor brochure in in-patient and out-patient settings. The training program is individually tailored according to their brunnstrom stage of the upper limb and hand, passive range of motion (PROM), muscle tone, muscle strength, fine motor abilities, grip strength and activities of daily living (ADL). It contains four parts of active exercises: exercises to improve and maintain joint mobility, therapeutic activities, functional training and activities of daily living training.

**Figure 3:**
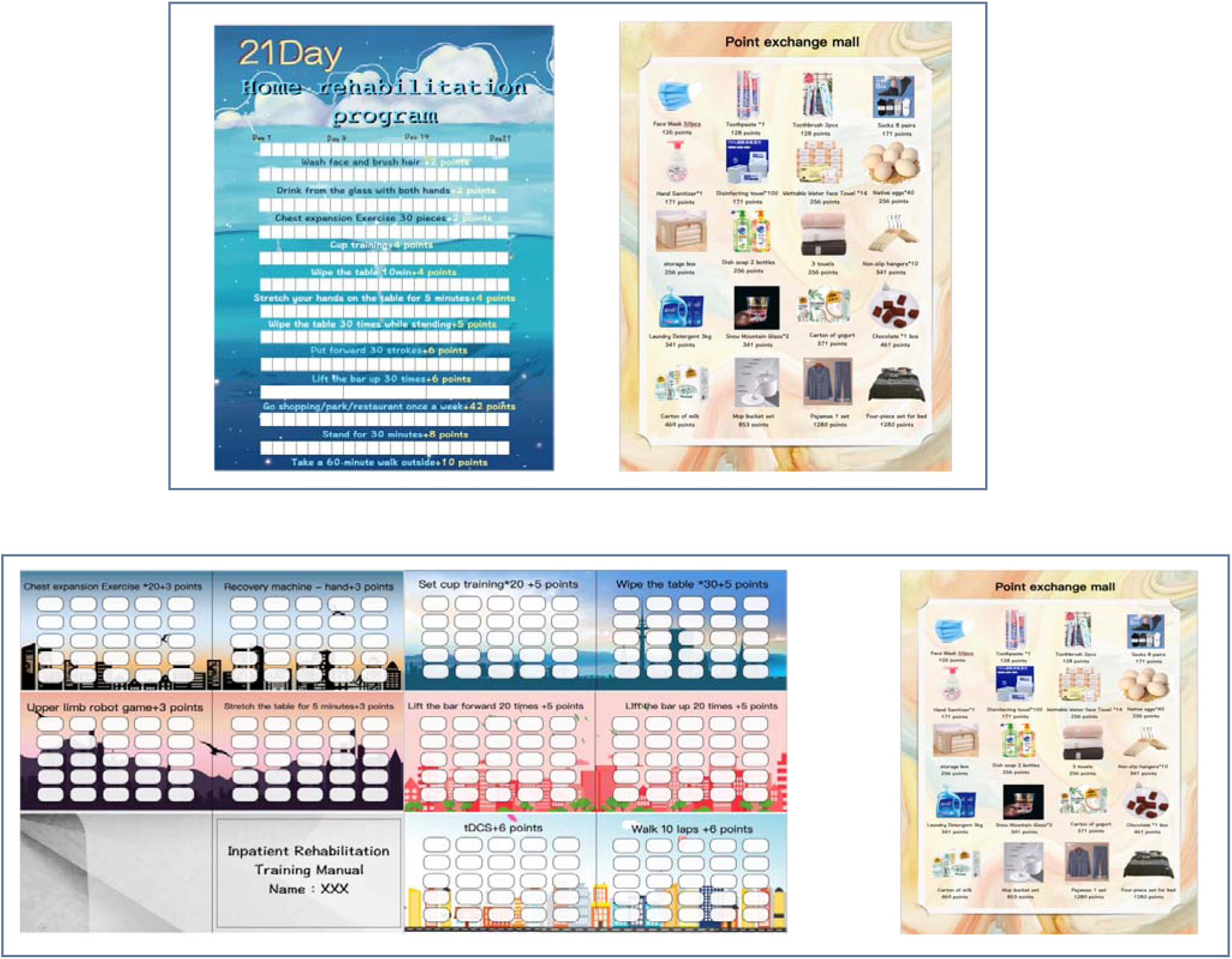
Schematic diagram of reward strategy self-monitor brochure in in-patient and out-patient settings

For RStDCS group and RS group, during the 21 day hospital stay, after a 20-minute tDCS stimulation, patients implement the program through self-report and free choice. The therapists will give the patient certain points according to the tasks completed by the patient every day. In the absence of supervision at home, the 21-day home program is designed for patients to choose their own task training and self-recording to monitor their intrinsic motivation. At the end of the plan, the points will be counted and exchanged for corresponding gifts (maximum available points can be 2142 which can exchange gifts worth 267 RMB).

## Outcomes measures

### Primary outcome measures

During the trial, the subjects will be assessed at 0, 3, 6 weeks and followed up at 3 months by means of assessment scales and behavioral observations. Basic subject data (gender, age, course of disease, etc.) will be obtained from the electronic medical record. All data will be entered into the paper Case Report Form (CRF) and uploaded to a study folder on our protected research server online.

Prior to the assessment, the assessor will be trained. Assessment scales include RMS (Rehabilitation Motivation Scale), FMA (Fugle-Meyer Assessment Scale), FIM (Function Independent Measure), ICF (ICF Activity and Participation Scale). RMS is designed by 1961 Litman to observe the motivation of patients to participate in physical therapy, and it was translated into Chinese version after discussion and modification by Professor Naiwen Guo’s team. The correlation coefficient of interrater reliability was 0.648 (p<0.001)^22^.

### Secondary outcome measures

FMA is a quantitative scale designed by Swedish doctor Fugle-Meyer on the basis of Brunnstrom rating Scale in 1980. At present, it is a relatively recognized and most widely used evaluation instrument, which has been proved to have good reliability and validity ^45^. FIM was proposed by researchers from the Functional Assessment Research Center of New York State in 1987. FIM more comprehensively and objectively reflects the ADL ability of stroke patients ^46^. The ICF Activity and Participation Scale evaluates the content of each item in six areas: understanding and communication, physical activity, self-care, getting along with others, life activity and social participation. It is applicable to the evaluation of the health status and health-related conditions of stroke population in the past 30 days. Behavioral observation data included the number of active exercise sessions supervised by therapists during hospitalization and the number of self-reported active exercise sessions by patients at home. After 42 days of reward training, the researchers collected the program manual and counted the number of completed sessions.

### Data management

Patient data will be collected using CRFS that meet the criteria of the GCP. The researchers will store the evaluation results in SPSS and back up the research folders on the protected research server regularly (every 3 months). Informed consent and CRF paper forms will be stored in a locked room in the institute’s office. All paper raw data and analysis procedures will be archived for subsequent researchers to reproduce or reuse the experimental data.

The data store uses the serial number as a marker to record the study results. The serial number key is kept by the principal investigator. Only authorized researchers can use the key during the study period, but not after the study is over. No participant information will be made public.

## Statistics

### Sample size

According to pretest, the efficacy validity of RStDCS on the rehabilitation motivation of post-stroke patients is 0.35. Using G*power software(3.1.9), Power=0.90, α=0.05; It is calculated that the minimum required sample size is 24 subjects in each group, 72 subjects in total in the three groups, and considering the 15% shedding rate, it is calculated that 29 subjects need to be included in each group, 87 subjects in total in the three groups.

### Statistical analyses

SPSS 26.0 version will be used for data analysis. Independent sample t test will be used to analyze the difference of active exercise frequency in chronic stroke patients with upper limb dysfunction with low motivation during hospitalization and at home after different treatments. In order to investigate the effect of rewarding training and neuroregulatory techniques on upper limb dysfunction of chronic stroke patients with low motivation over time, with rehabilitation motivation, motor function, functional independence, ICF activity and participation as dependent variables, 3 (groups: reward strategy combined with tDCS treatment(RStDCS) group/ rewarding strategy with sham tDCS treatment group / conventional treatment) × 3 (test type: 0 weeks /3 weeks /6 weeks) repeated measure ANOVA will be performed. For repeated measurement data, no data loss can occur due to the strict requirement. If the sample lacks measurement data, all the sample data should be removed from the model during the analysis.

p value less than 0.05, two-tailed, was considered statistically significant.

## Ethics And dissemination

### Ethics

Ethics approval was granted by the Ethics Committee of Yueyang Hospital of Integrated Chinese and Western Medicine Affiliated to Shanghai University of Traditional Chinese Medicine (2021-122). All participating patients will provide written informed consent.

All adverse events reported by the subject or discovered by the investigator will be recorded. The process of the incident and the analysis report will also be recorded. The following complications may be adverse events of Interest (AEIs) according to the previous research literature and the process of this study: Itchy, red and burning skin, headache, tingling, fatigue, manic or hypomanic episodes, seizures. All adverse events will be reported to the project manager, applicant and clinical trial institution.

### Safety

According to the guidelines for ethical review of drug clinical trials issued by the State Food and Drug Administration in 2010, the Ethics Committee of Yueyang Hospital of Integrated Traditional Chinese and Western Medicine Affiliated to Shanghai University of Traditional Chinese Medicine will assign a staff independent from the research to conduct an annual follow-up review according to the regulations, including the progress of the experiment, the number of subjects included, the number of completed cases, the number of withdrawn cases, the treatment of serious adverse events and any events or new information that may affect the study.

Amendment refers to the any modification to the test protocol during the course of the test. Any modification to the test plan during the test shall be submitted to the Ethics Committee for review and approval before implementation. The Ethics Committee shall request sponsors and/or researchers to submit information related to the amendment review, including (but not limited to): Contents of the amendment and reasons for the amendment; the impact of the modification plan on the expected risks and benefits; the impact of the modification plan on the rights and safety of the subjects.

### Dissemination

Authorship eligibility will be assured in accordance with The International Committee of Medical Journal Editors (ICMJE) guidelines. The results will be disseminated through (open access) peer-reviewed publications, networks of scientists, patient associations, professionals and the public, and presented at relevant conferences. Participants of the study will be updated about the progress and results of the study by newsletters.

## Discussion

This randomized controlled trial was designed to investigate the efficacy of RstDCS and Rs in improving patients’ motivation compared to traditional rehabilitation therapy in 87 patients. The safety of RstDCS was observed and its effects on motor function, ability of daily living, ICF activity and social participation were compared.

## Data Availability

All data produced in the present study are available upon reasonable request to the authors

## Acknowledgements

We thank patients for their commitment and participation in this trial; We would like to thank the doctors and therapists in the Rehabilitation Department of Yueyang Hospital of Integrated Traditional Chinese and Western Medicine of Shanghai University of Traditional Chinese Medicine for their help in patient recruitment and rehabilitation program design. Thanks to Dong sheng Xu and Xia Shen for their help in the design and content of the study as well as the funding of the project.

## Supplementary materials

**Supplementary File 1:** Rehabilitation Motivation Scale

Rehabilitation Motivation Scale
|  | 1 | 2 | 3 | 4 |
| --- | --- | --- | --- | --- |
| Does the patient want to do rehabilitation exercises? | never | few | often | always |
| Does the patient complain of poor recovery? | never | few | often | always |
| Does the patient complain of excessive pain? | never | few | often | always |
| Will the patient try to do rehabilitation exercises? | never | few | often | always |
| Does the patient need too much encouragement? | never | few | often | always |
| Does the patient cooperate? | never | few | often | always |
| Does the patient find excuses to avoid recovery? | never | few | often | always |
| Does the patient ask or seek for resources? | never | few | often | always |

**Supplementary File 2:**
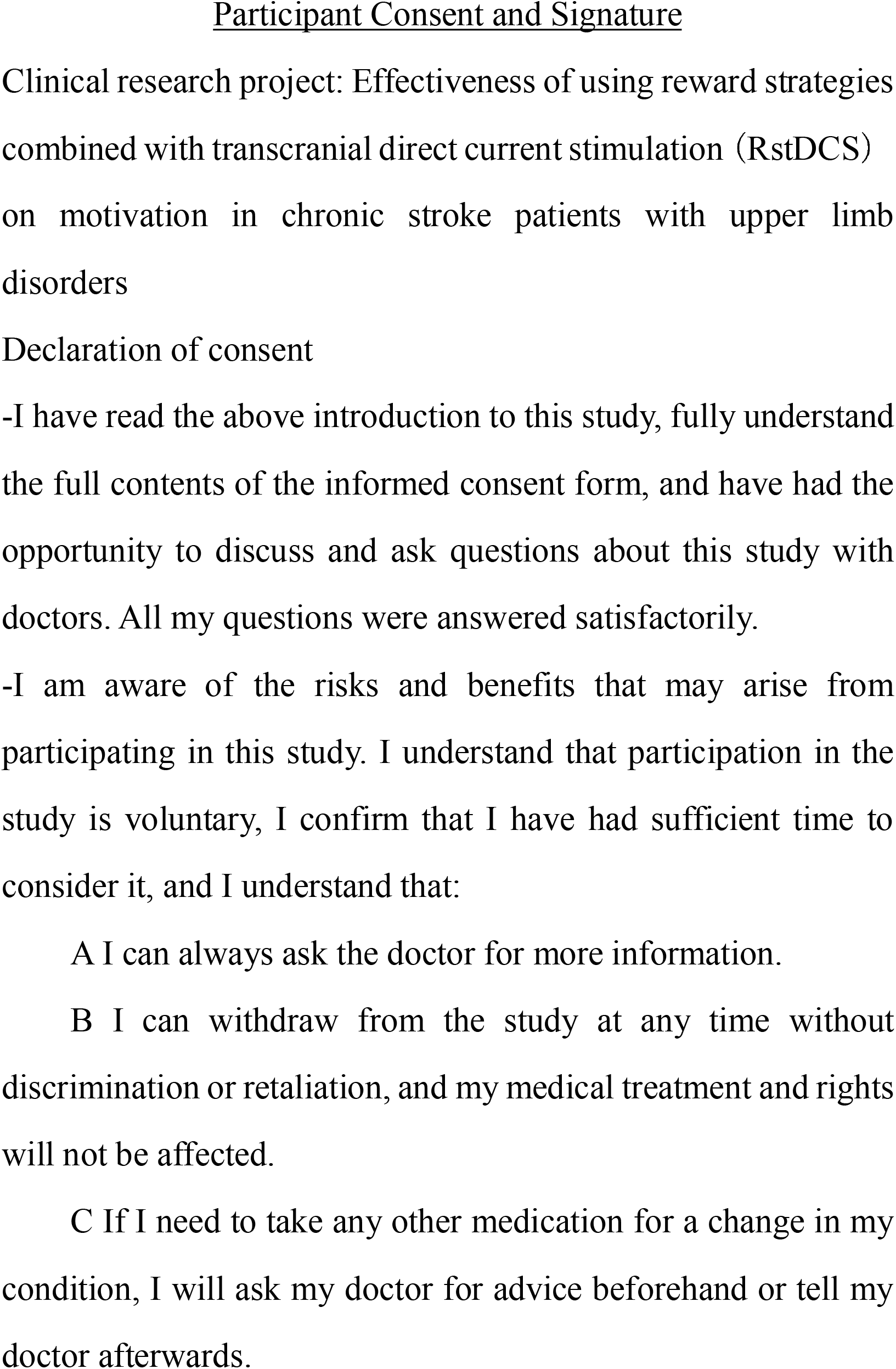
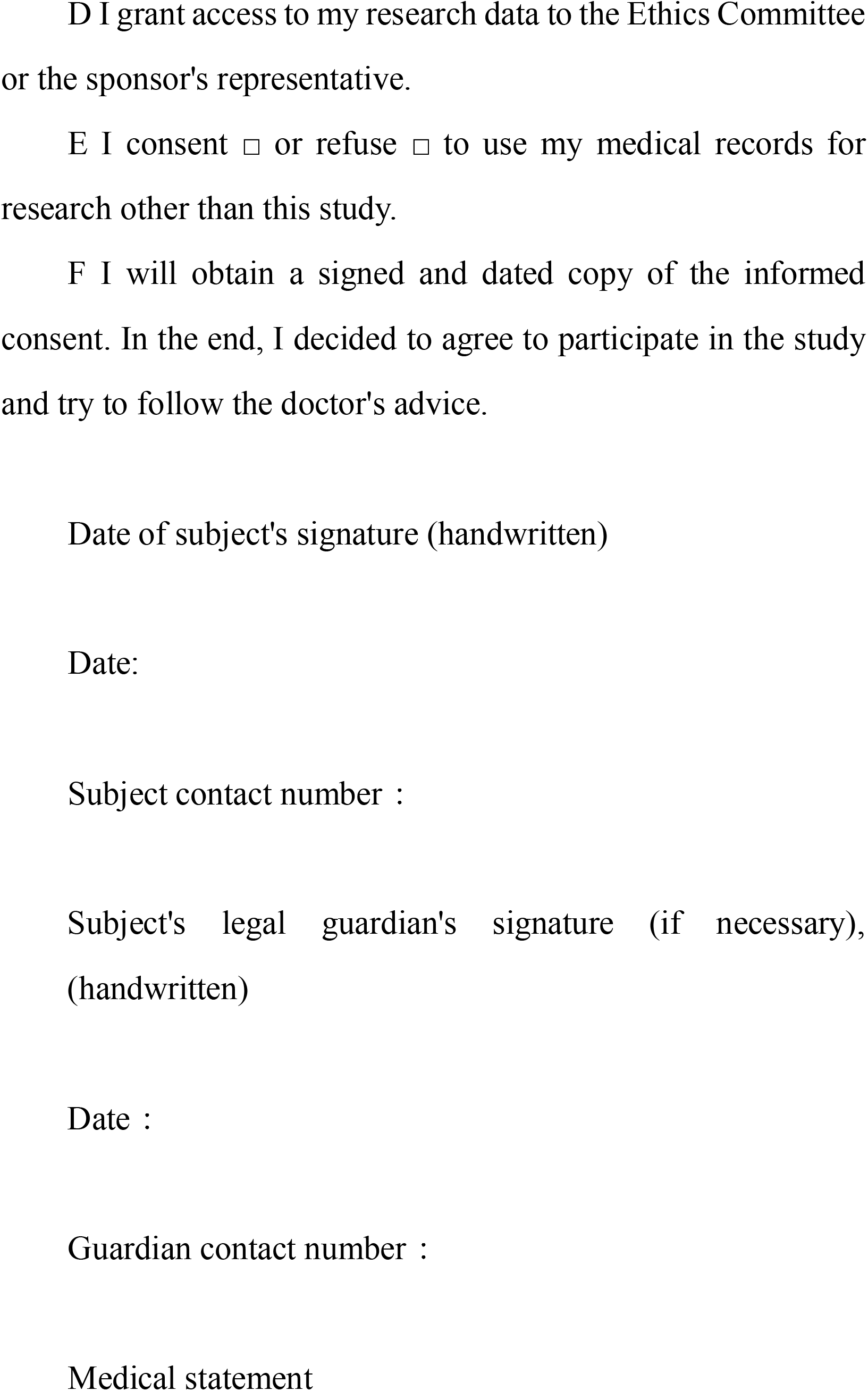
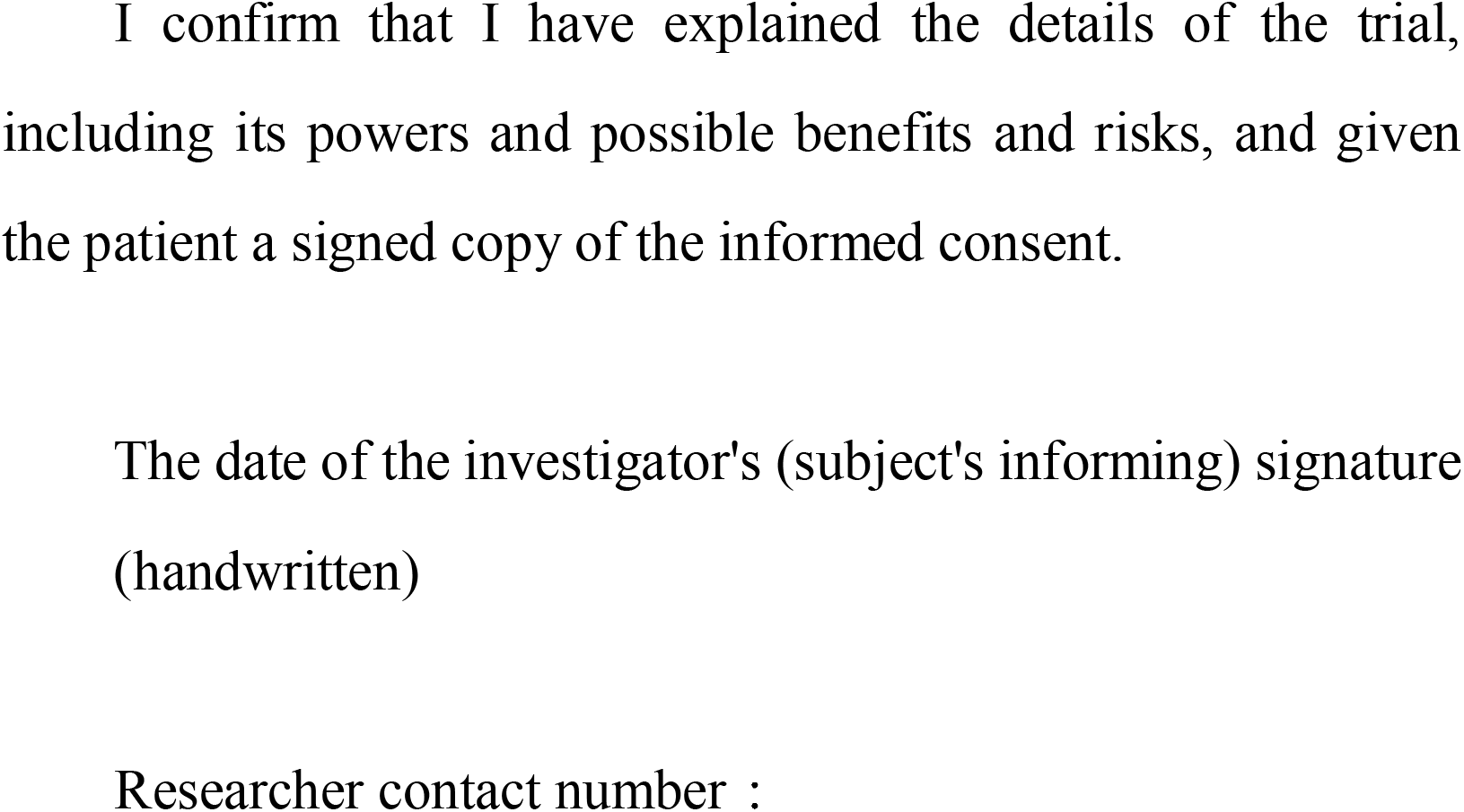
Consent Form Signed by participants

## Footnotes

### Author Contributions

PZ: Proposal and protocol development, acquiring of data, trial coordination. WL: Optimizing laboratory protocols, acquiring of data, trial coordination. SC, WZ and YC: Protocol development, optimizing the research protocols. DX: Principal Investigator. XS: Principal Investigator. All authors read and approved the final manuscript.

## Funding

This project was funded by the Key project of the National Ministry of Science and Technology "Neuroregulation and limb movement synergistic Enhancement rehabilitation Technology Research" (2020YFC2004202). The project was jointly supported by Tongji University, Yueyang Hospital of Integrated Traditional Chinese and Western Medicine Affiliated to Shanghai University of Traditional Chinese Medicine and Shuguang Hospital Affiliated to Shanghai University of Traditional Chinese Medicine. The project funders was not involved in the collection, analysis and interpretation of the data, nor in the writing of the manuscript.

## Patient and public involvement

This study involved working with patients and their families to develop personalized home and inpatient rehabilitation plans. The joint development of rehabilitation goals and plans could improve patients’ motivation to recover, so this was also part of the experiment, and also expanded the active exercise task set. Throughout the study, researchers will interview patients about their feelings, satisfaction and dissatisfaction during the whole study process, and methods that need to be improved, so as to improve the paradigm in future studies.

## Data Availability Statement

The data set of this study can be obtained with the permission of the corresponding author and reasonable requirements, and the data acquisition must comply with the rules of research data acquisition of Tongji University and Shanghai University of Traditional Chinese Medicine.

## Competing interests

The authors declare that they have no competing interests.

## Patient consent for publication

Not required.

## Provenance and peer review

Not commissioned;externally peer reviewed.

## Reporting checklist for protocol of a clinical trial

Based on the SPIRIT guidelines.

### Instructions to authors

Complete this checklist by entering the page numbers from your manuscript where readers will find each of the items listed below.

Your article may not currently address all the items on the checklist. Please modify your text to include the missing information. If you are certain that an item does not apply, please write "n/a" and provide a short explanation.

Upload your completed checklist as an extra file when you submit to a journal.

In your methods section, say that you used the SPIRITreporting guidelines, and cite them as:

Chan A-W, Tetzlaff JM, Gøtzsche PC, Altman DG, Mann H, Berlin J, Dickersin K, Hróbjartsson A, Schulz KF, Parulekar WR, Krleža-Jerić K, Laupacis A, Moher D. SPIRIT 2013 Explanation and Elaboration: Guidance for protocols of clinical trials. BMJ. 2013;346:e7586

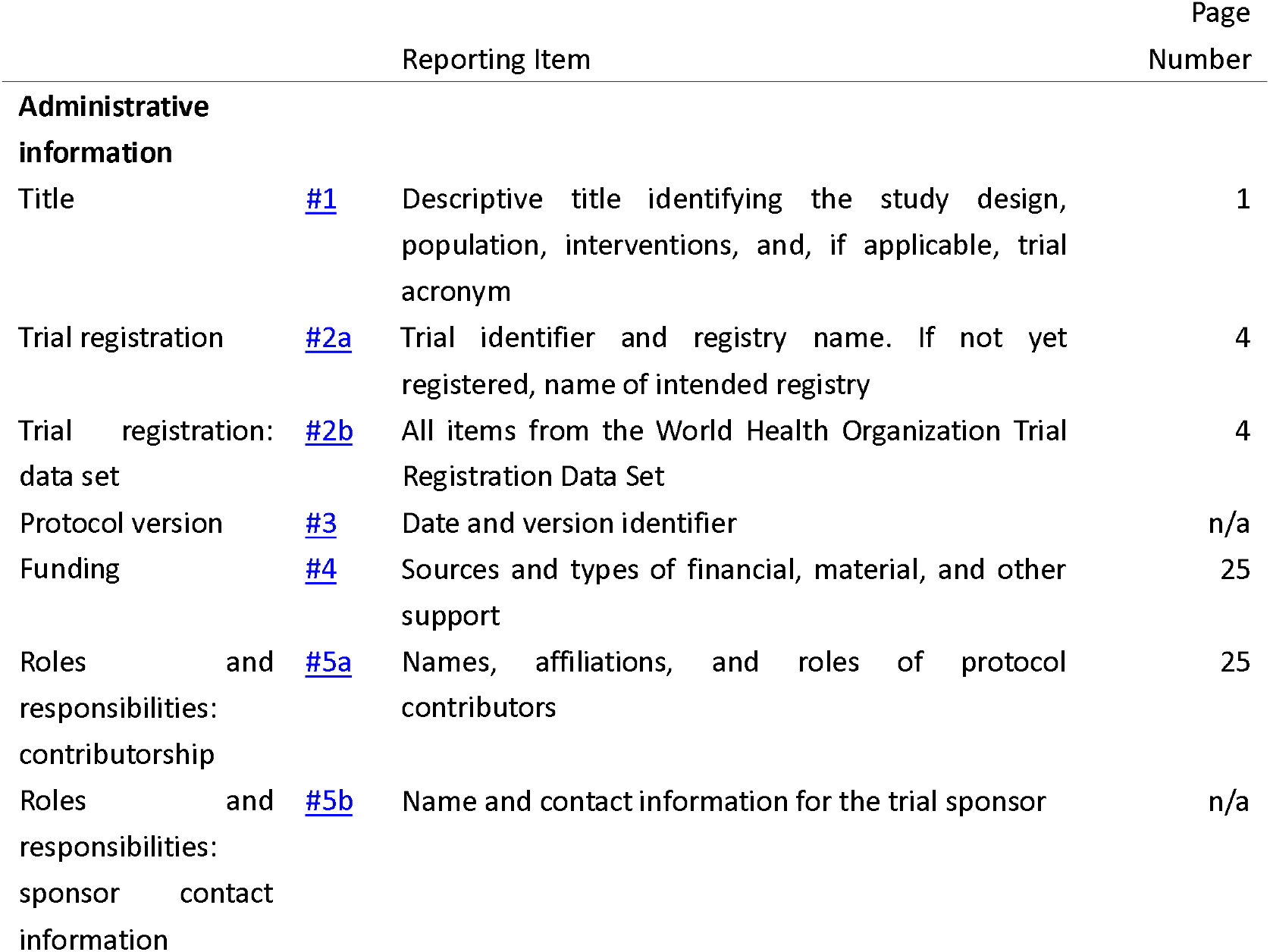

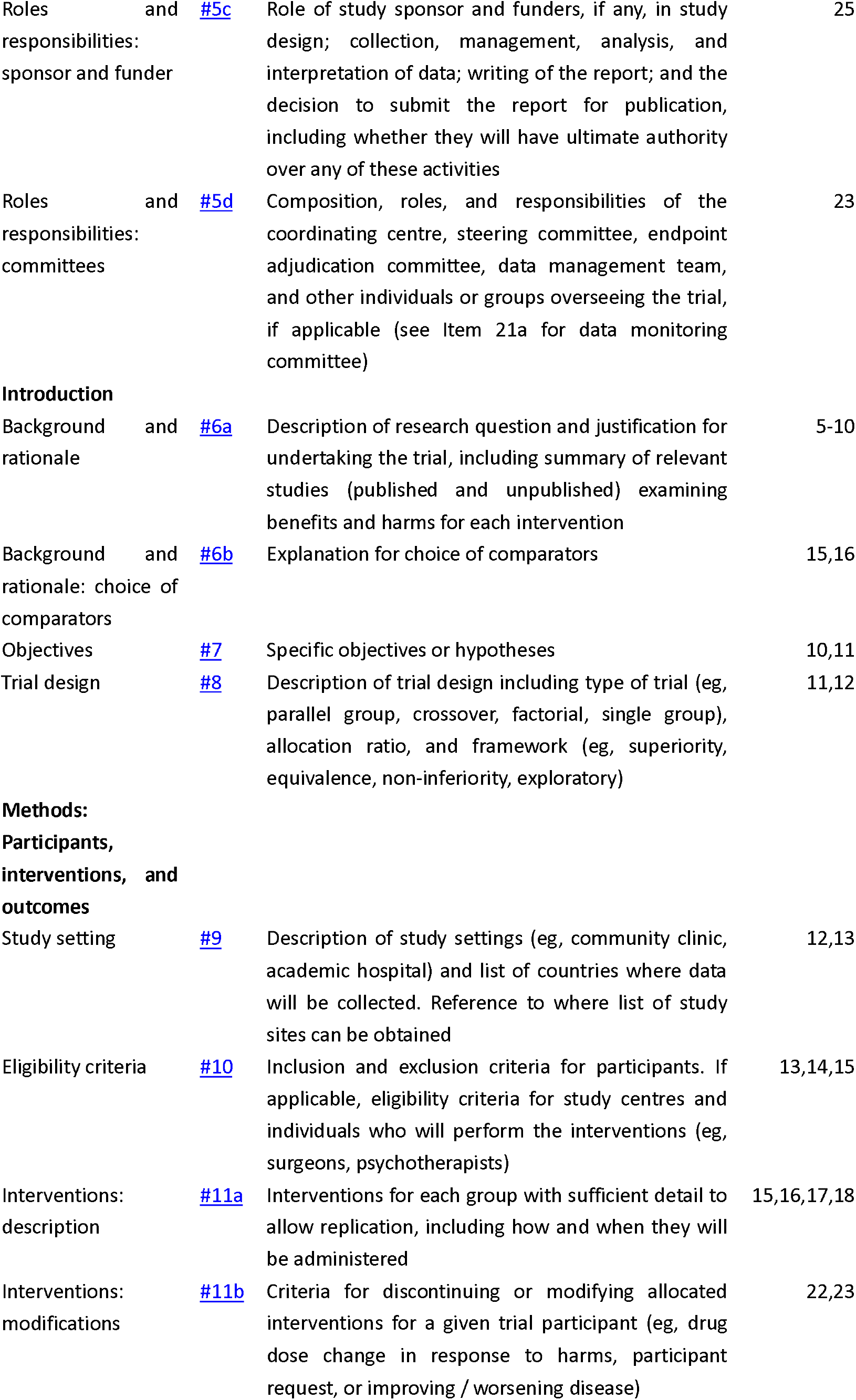

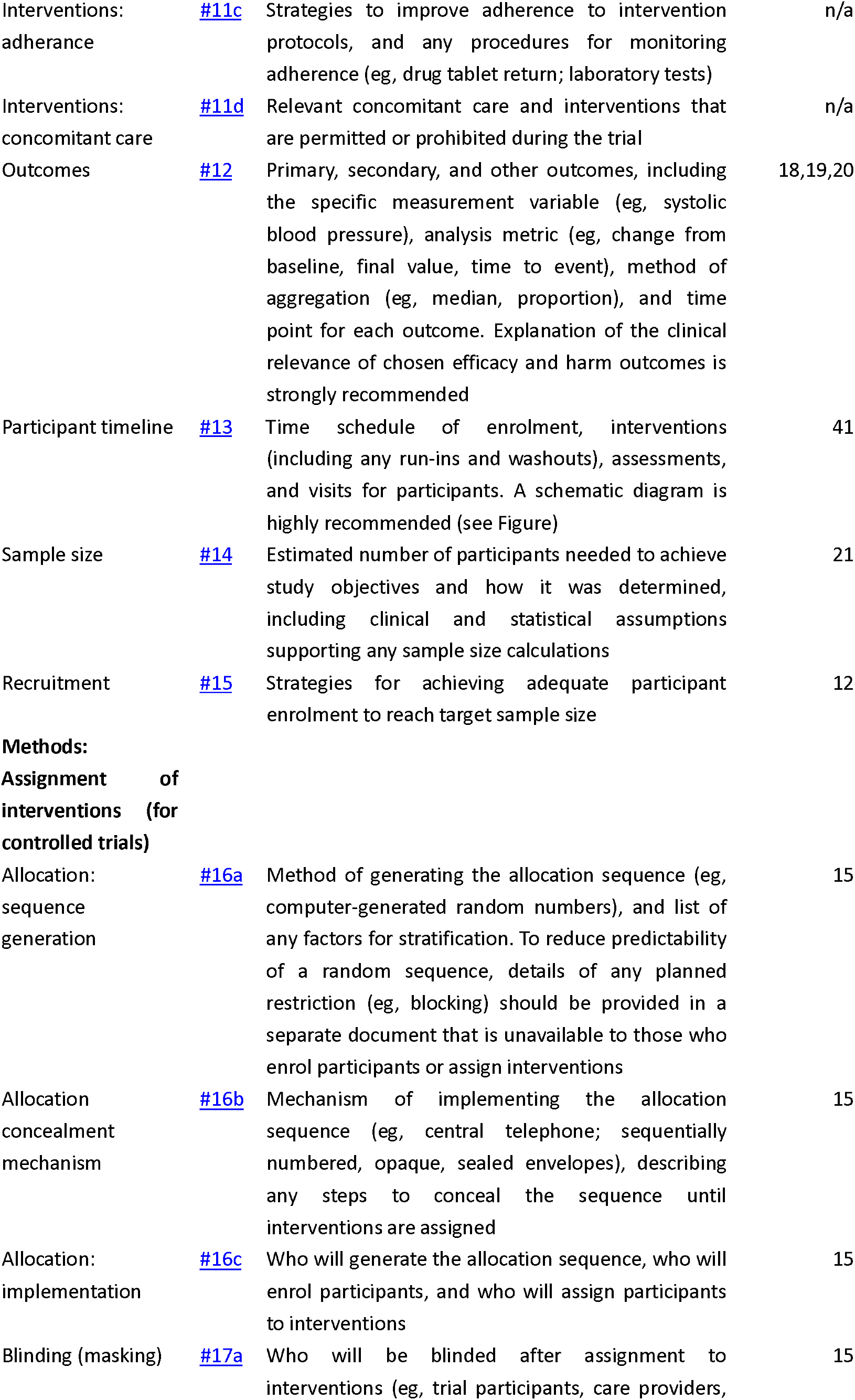

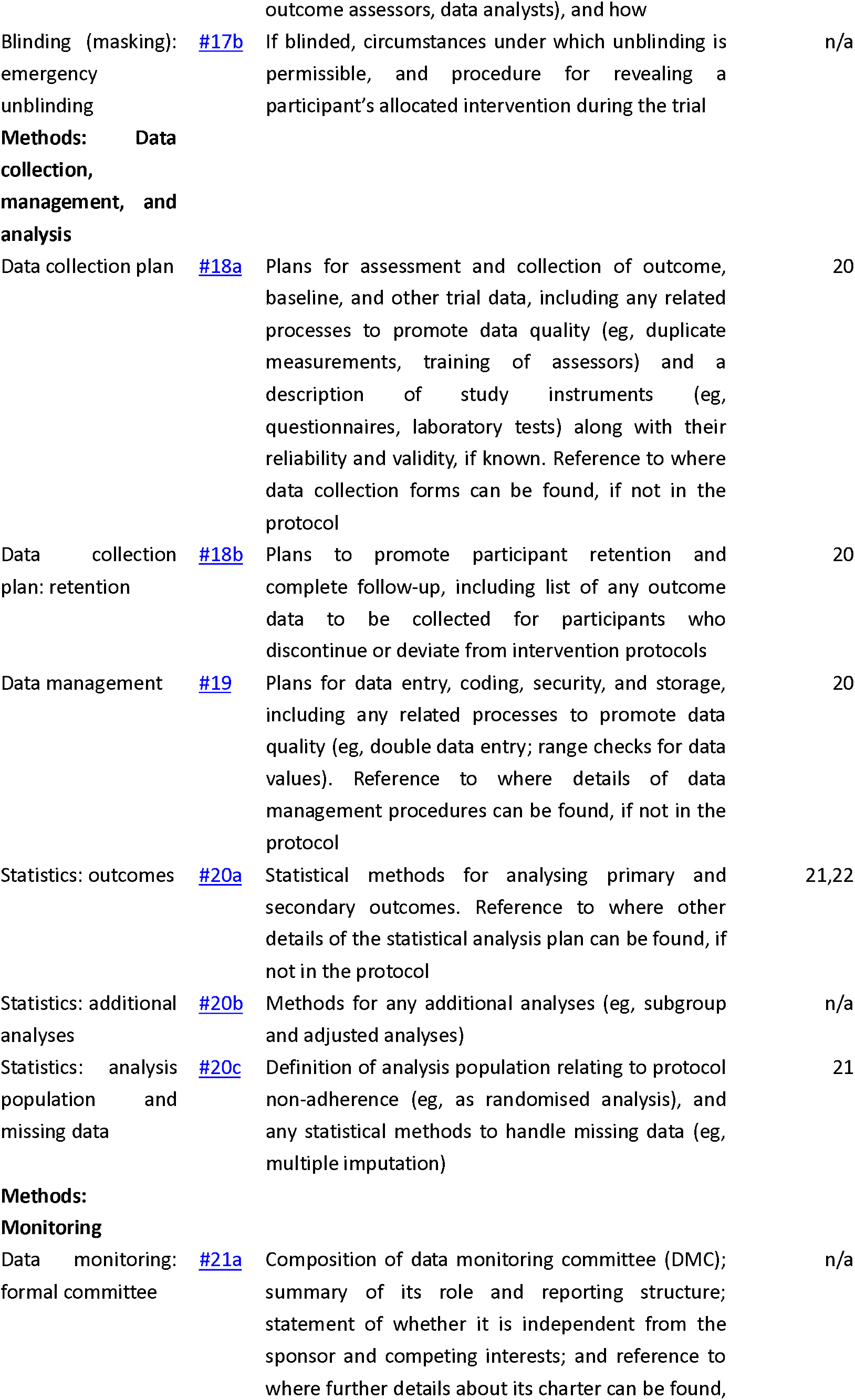

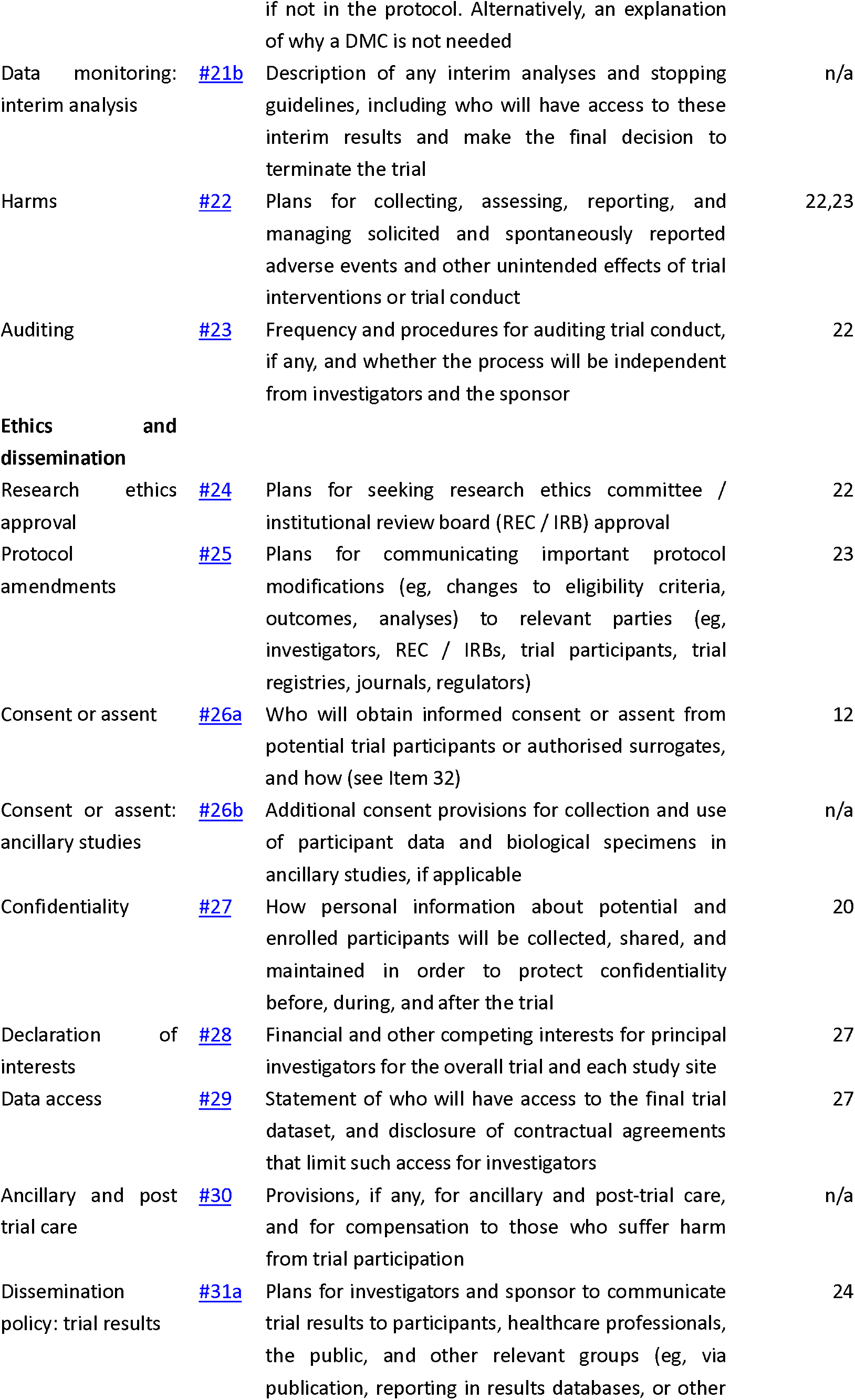

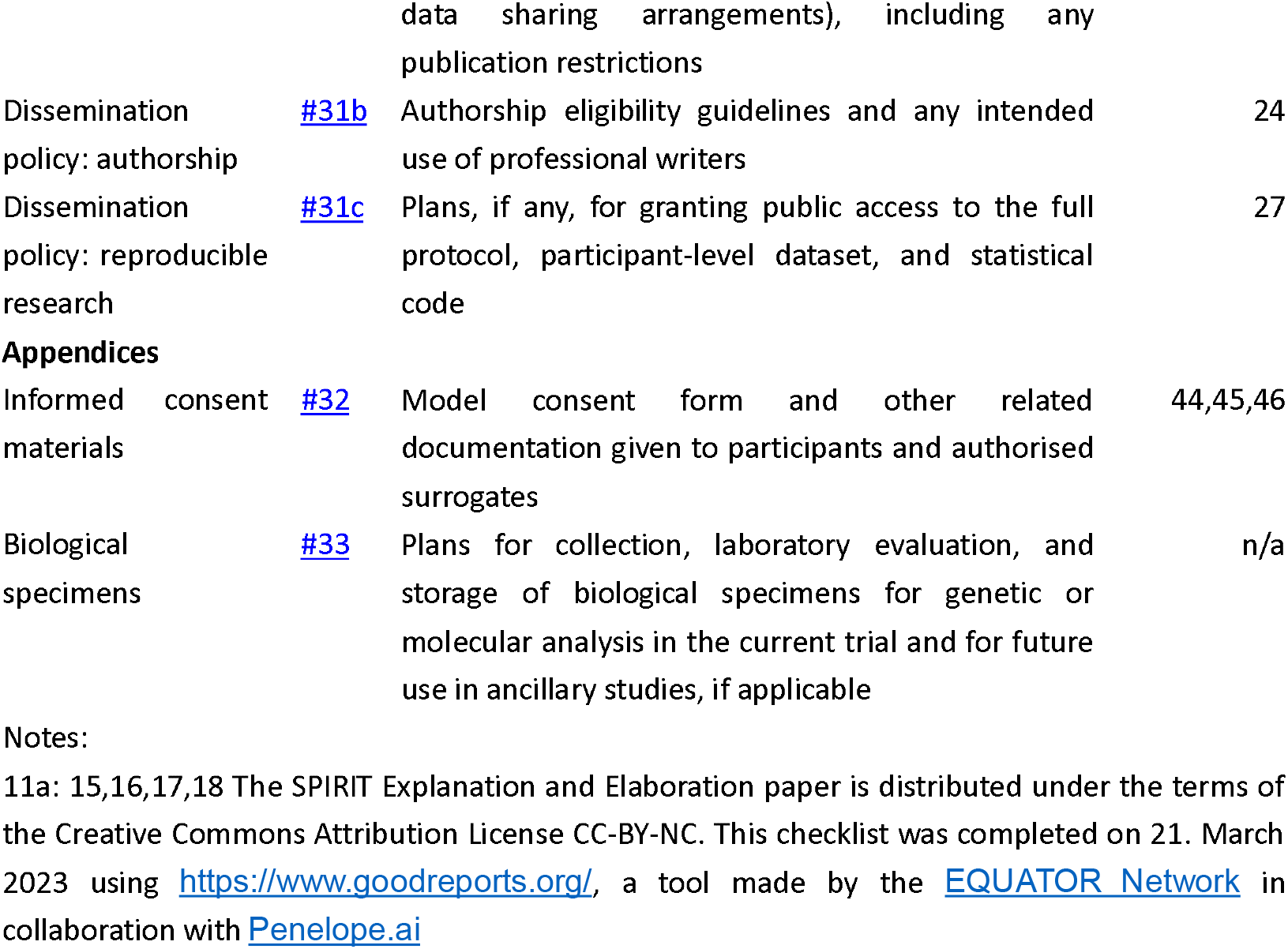

## Notes

### Competing Interest Statement

The authors have declared no competing interest.

### Clinical Trial

ChiCTR2300069068

### Author Declarations

the Ethics Committee of Yueyang Hospital of Integrated Chinese and Western Medicine Affiliated to SHUTCM gave ethical approval for this work

## References

1. G. Broeks, J, Lankhorst, GJ, Rumping, K, Prevo AJH. The long-term outcome of arm function after stroke: results of a follow-up study. Disability and Rehabilitation. 1999;21(8):357–364. doi:10.1080/096382899297459

2. Chen L, Xiong S, Liu Y, et al. Comparison of Motor Relearning Program versus Bobath Approach for Prevention of Poststroke Apathy: A Randomized Controlled Trial. Journal of Stroke and Cerebrovascular Diseases. 2019;28(3):655–664. doi:10.1016/j.jstrokecerebrovasdis.2018.11.011

3. Lohse KR, Lang CE, Boyd LA. Is More Better? Using Metadata to Explore Dose–Response Relationships in Stroke Rehabilitation. Stroke. 2014;45(7):2053–2058. doi:10.1161/STROKEAHA.114.004695

4. Olgiati E, Russell C, Soto D, Malhotra P. Motivation and attention following hemispheric stroke. In: Progress in Brain Research. Vol 229. Elsevier; 2016:343-366. doi:10.1016/bs.pbr.2016.06.011

5. Broderick M, Almedom L, Burdet E, Burridge J, Bentley P. Self-Directed Exergaming for Stroke Upper Limb Impairment Increases Exercise Dose Compared to Standard Care. Neurorehabil Neural Repair. 2021;35(11):974–985. doi:10.1177/15459683211041313

6. Maclean N, Pound P, Wolfe C, Rudd A. The Concept of Patient Motivation: A Qualitative Analysis of Stroke Professionals’ Attitudes. Stroke. 2002;33(2):444–448. doi:10.1161/hs0202.102367

7. Cappon D, den Boer T, Jordan C, et al. Safety and Feasibility of Tele-Supervised Home-Based Transcranial Direct Current Stimulation for Major Depressive Disorder. Front Aging Neurosci. 2022;13:765370. doi:10.3389/fnagi.2021.765370

8. Apathy after stroke: Diagnosis, mechanisms, consequences, and treatment - Jonathan Tay, Robin G Morris, Hugh S Markus, 2021. Accessed December 4, 2022. https://journals.sagepub.com/doi/10.1177/1747493021990906

9. Mikami K, Jorge RE, Moser DJ, Jang M, Robinson RG. Incident Apathy During the First Year After Stroke and Its Effect on Physical and Cognitive Recovery. The American Journal of Geriatric Psychiatry. 2013;21(9):848–854. doi:10.1016/j.jagp.2013.03.012

10. Douven E, Köhler S, Rodriguez MMF, Staals J, Verhey FRJ, Aalten P. Imaging Markers of Post-Stroke Depression and Apathy: a Systematic Review and Meta-Analysis. Neuropsychol Rev. 2017;27(3):202–219. doi:10.1007/s11065-017-9356-2

11. Yang S, Hua P, Shang X, et al. Deficiency of brain structural sub-network underlying post-ischaemic stroke apathy. Eur J Neurol. 2015;22(2):341–347. doi:10.1111/ene.12575

12. Le Heron C, Apps. MAJ, Husain M. The anatomy of apathy: A neurocognitive framework for amotivated behaviour. Neuropsychologia. 2018;118:54–67. doi:10.1016/j.neuropsychologia.2017.07.003

13. Rochat L, Van der Linden M, Renaud O, et al. Poor reward sensitivity and apathy after stroke: Implication of basal ganglia. Neurology. 2013;81(19):1674–1680. doi:10.1212/01.wnl.0000435290.49598.1d

14. Gual N, Pérez LM, Castellano-Tejedor C, et al. IMAGINE study protocol of a clinical trial: a multi-center, investigator-blinded, randomized, 36-month, parallel-group to compare the effectiveness of motivational interview in rehabilitation of older stroke survivors. BMC Geriatr. 2020;20(1):321. doi:10.1186/s12877-020-01694-6

15. Fletcher S, Kulnik ST, Demain S, Jones F. The problem with self-management: Problematising self-management and power using a Foucauldian lens in the context of stroke care and rehabilitation. Collins SM, ed. PLoS ONE. 2019;14(6):e0218517. doi:10.1371/journal.pone.0218517

16. Ostir GV, Berges IM, Ottenbacher ME, Clow A, Ottenbacher KJ. Associations Between Positive Emotion and Recovery of Functional Status Following Stroke. Psychosomatic Medicine. 2008;70(4):404–409. doi:10.1097/PSY.0b013e31816fd7d0

17. Watkins CL, Wathan JV, Leathley MJ, et al. The 12-Month Effects of Early Motivational Interviewing After Acute Stroke: A Randomized Controlled Trial. Stroke. 2011;42(7):1956–1961. doi:10.1161/STROKEAHA.110.602227

18. Tay J, Tuladhar AM, Hollocks MJ, et al. Apathy is associated with large-scale white matter network disruption in small vessel disease. Neurology. Published online February 8, 2019:10.1212/WNL.0000000000007095. doi:10.1212/WNL.0000000000007095

19. Jang JY, Han SD, Yew B, et al. Resting-State Functional Connectivity Signatures of Apathy in Community-Living Older Adults. Front Aging Neurosci. 2021;13:691710. doi:10.3389/fnagi.2021.691710

20. Chong TTJ, Husain M. The role of dopamine in the pathophysiology and treatment of apathy. In: Progress in Brain Research. Vol 229. Elsevier; 2016:389-426. doi:10.1016/bs.pbr.2016.05.007

21. Quattrocchi G, Bestmann S. Journal Club: Possible role of the basal ganglia in poor reward sensitivity and apathy after stroke. Neurology. 2014;82(20):e171–e173. doi:10.1212/WNL.0000000000000439

22. Chen HM, Lee HL, Yang FC, Chiu YW, Chao SY. Effectiveness of Motivational Interviewing in Regard to Activities of Daily Living and Motivation for Rehabilitation among Stroke Patients. IJERPH. 2020;17(8):2755. doi:10.3390/ijerph17082755

23. Qiqi N, Hangting L, Jia W, Jiaoni S, Xinrui W, Guijuan H. A Meta-analysis of the Effect of Motivational Interviewing on Depression, Anxiety, and Quality of Life in Stroke Patients. Journal of Neuroscience Nursing. 2021;53(6):244–250. doi:10.1097/JNN.0000000000000617

24. Fishman KN, Ashbaugh AR, Swartz RH. Goal Setting Improves Cognitive Performance in a Randomized Trial of Chronic Stroke Survivors. Stroke. 2021;52(2):458–470. doi:10.1161/STROKEAHA.120.032131

25. Dörfler E, Kulnik ST. Despite communication and cognitive impairment – person-centred goal-setting after stroke: a qualitative study. Disability and Rehabilitation. 2020;42(25):3628–3637. doi:10.1080/09638288.2019.1604821

26. Scobbie L, Brady MC, Duncan EAS, Wyke S. Goal attainment, adjustment and disengagement in the first year after stroke: A qualitative study. Neuropsychological Rehabilitation. 2021;31(5):691–709. doi:10.1080/09602011.2020.1724803

27. Signal N, McPherson K, Lewis G, et al. What influences acceptability and engagement with a high intensity exercise programme for people with stroke? A qualitative descriptive study. Danzl M, Etter N, eds. NRE. 2016;39(4):507–517. doi:10.3233/NRE-161382

28. Studer B, Timm A, Sahakian BJ, Kalenscher T, Knecht S. A decision-neuroscientific intervention to improve cognitive recovery after stroke. Brain. 2021;144(6):1764–1773. doi:10.1093/brain/awab128

29. Waddell KJ, Patel MS, Clark K, Harrington TO, Greysen SR. Leveraging insights from behavioral economics to improve mobility for adults with stroke: Design and rationale of the BE Mobile clinical trial. Contemporary Clinical Trials. 2021;107:106483. doi:10.1016/j.cct.2021.106483

30. Bandura A. Social Cognitive Theory: An Agentic Perspective. Annu Rev Psychol. 2001;52(1):1–26. doi:10.1146/annurev.psych.52.1.1

31. Ryan RM, Deci EL. Self-determination theory and the facilitation of intrinsic motivation, social development, and well-being. American Psychologist. 2000;55(1):68–78. doi:10.1037/0003-066X.55.1.68

32. Deci EL, Ryan RM. Self-determination theory in health care and its relations to motivational interviewing: a few comments. Int J Behav Nutr Phys Act. 2012;9(1):24. doi:10.1186/1479-5868-9-24

33. Timmermans AA, Seelen HA, Willmann RD, Kingma H. Technology-assisted training of arm-hand skills in stroke: concepts on reacquisition of motor control and therapist guidelines for rehabilitation technology design. J NeuroEngineering Rehabil. 2009;6(1):1. doi:10.1186/1743-0003-6-1

34. Timmermans AAA, Seelen HAM, Geers RPJ, et al. Sensor-Based Arm Skill Training in Chronic Stroke Patients: Results on Treatment Outcome, Patient Motivation, and System Usability. IEEE Trans Neural Syst Rehabil Eng. 2010;18(3):284–292. doi:10.1109/TNSRE.2010.2047608

35. Finley M, Combs S. User perceptions of gaming interventions for improving upper extremity motor function in persons with chronic stroke. Physiotherapy Theory and Practice. 2013;29(3):195–201. doi:10.3109/09593985.2012.717591

36. Johnson MJ, Shakya Y, Strachota E, Ahamed SI. Low-cost monitoring of patients during unsupervised robot/computer assisted motivating stroke rehabilitation. Biomedizinische Technik/Biomedical Engineering. 2011;56(1):5–9. doi:10.1515/bmt.2010.050

37. Leow L, Marinovic W, Rugy A, Carroll TJ. Task errors contribute to implicit aftereffects in sensorimotor adaptation. Eur J Neurosci. 2018;48(11):3397–3409. doi:10.1111/ejn.14213

38. Mitaki S, Onoda K, Abe S, Oguro H, Yamaguchi S. The Effectiveness of Repetitive Transcranial Magnetic Stimulation for Poststroke Apathy Is Associated with Improved Interhemispheric Functional Connectivity. Journal of Stroke and Cerebrovascular Diseases. 2016;25(12):e219–e221. doi:10.1016/j.jstrokecerebrovasdis.2016.05.014

39. Widmer M, Lutz K, Luft AR. Reduced striatal activation in response to rewarding motor performance feedback after stroke. NeuroImage: Clinical. 2019;24:102036. doi:10.1016/j.nicl.2019.102036

40. Manuel AL, Murray NWG, Piguet O. Transcranial direct current stimulation (tDCS) over vmPFC modulates interactions between reward and emotion in delay discounting. Sci Rep. 2019;9(1):18735. doi:10.1038/s41598-019-55157-z

41. Kelley NJ, Gallucci A, Riva P, Romero Lauro LJ, Schmeichel BJ. Stimulating Self-Regulation: A Review of Non-invasive Brain Stimulation Studies of Goal-Directed Behavior. Front Behav Neurosci. 2019;12:337. doi:10.3389/fnbeh.2018.00337

42. Wang H, Yu H, Liu M, et al. Effects of tDCS on Brain Functional Network of Patients After Stroke. IEEE Access. 2020;8:205625–205634. doi:10.1109/ACCESS.2020.3037924

43. Meng L, Ma Q. Live as we choose: The role of autonomy support in facilitating intrinsic motivation. International Journal of Psychophysiology. 2015;98(3):441–447. doi:10.1016/j.ijpsycho.2015.08.009

44. Soutschek A, Tobler PN. Causal role of lateral prefrontal cortex in mental effort and fatigue. Hum Brain Mapp. 2020;41(16):4630–4640. doi:10.1002/hbm.25146

45. Hsieh YW, Hsueh IP, Chou YT, Sheu CF, Hsieh CL, Kwakkel G. Development and Validation of a Short Form of the Fugl-Meyer Motor Scale in Patients With Stroke. Stroke. 2007;38(11):3052–3054. doi:10.1161/STROKEAHA.107.490730

46. Heinemann AW, Michael Linacre J, Wright BD, Hamilton BB, Granger C. Measurement characteristics of the Functional Independence Measure. Topics in Stroke Rehabilitation. 1994;1(3):1–15. doi:10.1080/10749357.1994.11754030

